# Investigating Sex Bias in Machine Learning Research: A Systematic Review in Rheumatoid Arthritis

**DOI:** 10.1101/2024.10.15.24315519

**Authors:** Anahita Talwar, Shruti Turner, Claudia Maw, Georgina Quayle, Thomas N Watt, Sunir Gohil, Emma Duckworth, Coziana Ciurtin

## Abstract

Unchecked sex bias in machine learning (ML) algorithms used in healthcare can exacerbate disparities in care and treatment. We aimed to assess the acknowledgment and mitigation of sex bias within studies using supervised ML for improving clinical outcomes in Rheumatoid Arthritis (RA). For this systematic review, we searched PUBMED and EMBASE for original, English language studies published between 2018 to November 2023. We scored papers on whether the authors reported, attempted to mitigate or successfully mitigated the following types of bias: training data bias, test data bias, input variable bias, output variable bias, analysis bias, and assessed the quality of ML research in all papers. This study is registered on PROSPERO with ID CRD42023431754. We identified 52 papers to include in our review. All but one had a female skew in their study participants, yet 42 papers did not acknowledge any potential sex bias. Three papers assessed bias in model performance by sex disaggregating their results. One paper acknowledged potential sex bias in input variables, and six papers in their output variables, predominantly disease activity scores. No paper attempted to mitigate for any type of sex bias. The findings demonstrate the requirement for increased promotion of inclusive and equitable ML practices in healthcare.

## Introduction

Machine Learning (ML) algorithms, computational models and techniques that use patterns learnt from data to make useful predictions, hold immense potential for advancing precision medicine in healthcare. The applications of ML remain wide, including disease diagnosis, pattern or image recognition and classification, drug development, and prognostic prediction tools for clinical use^1^. The implementation of ML models into real-world settings raises ethical concerns that necessitate careful consideration during model development to produce fair algorithms^2–4^. ML algorithms learn from existing data, and if the data contains biases or imbalances, it can result in inequalities in the model’s predictions, ultimately affecting the model’s clinical utility and relevance for the wider population. Thus, ensuring the equity and fairness of developed algorithms when implemented into real-world settings is crucial, and commands the consideration of often overlooked individual differences in healthcare, including sex and gender disparities.

### Sex Bias in ML Research in Healthcare

There has been an increasing emphasis on sex and gender as variables in analysis in healthcare research^5–8^. This is in part driven by an evolving recognition of the substantial influence that sex and gender differences exert on symptomatology, disease phenotypes, treatment responses and experience of care^9–12^. Additionally, it stems from an increased awareness of the historical underrepresentation of women in clinical and preclinical trials, as well as in research professions^13,14^, and the consequential adverse implications for women in clinical practice^15,16^. When parameters related to, derived from, or significantly impacted by an individual’s sex, are successfully accounted for in model development, ML algorithms can contribute to improved equity in the quality of care for both sexes, such as AwareDx, developed to predict sex differences in drug response and in doing so, to minimise adverse events and provide tailored drug dosing^17^. When disregarded or unsuccessfully mitigated, bias can result in inequitable care and treatment for patients of either sex, and ultimately unfair outcomes.

A common source of bias arises when certain groups are underrepresented in the training and test data. Consequently, model predictions will suffer for such underrepresented groups^18,19^. Additionally, biases can originate from variables used as predictors in the model, such as healthcare measures that are more representative of symptoms experienced by one sex. In real world terms, this can exacerbate disparities in diagnosis, treatment decisions, and access to care, perpetuating inequities in healthcare. To address this issue proactively, it is essential to critically evaluate and refine ML algorithms, ensuring they are sensitive to and account for sex, as well as gender differences. By mitigating bias in algorithmic decision-making processes, healthcare systems can strive for more equitable and inclusive care delivery, ultimately improving health outcomes for all individuals.

Rheumatoid Arthritis (RA) is a strong candidate for exploring sex bias in ML due to its relatively high prevalence in the general population, female predominance irrespective of age at onset, as well as the potential for ML to facilitate precision medicine.

### Using RA to explore gender bias in ML

RA is a chronic autoimmune disease, with an estimated 17.8 million individuals living with it worldwide in 2020^20^, generally characterised by inflammation of the synovial joints, leading to pain, stiffness, and swelling, which severely impact mobility and quality of life. However, RA is a heterogeneous condition with individuals exhibiting diverse immunomolecular profiles and clinical phenotypes^21,22^, varying responses to established and emerging treatments^23–25^ and complexities in comorbidity management, despite clear benefits of prompt diagnosis and pharmacological intervention^26–30^. ML offers a promising contribution to optimal RA management by identifying complex patterns in patient data (e.g. reported symptoms, genetics, blood and imaging biomarkers, etc.), and improving predictions related to patient outcomes^31–34^. Finally, sex differences have been reported in RA, such as a three-fold increased prevalence in women versus men overall, poorer symptoms scores, greater impact on quality of life and increased prevalence of irreversible joint damage in women compared to men^35,36^, and reduced treatment efficacy and adherence in women^37–40^. There are several potential and interdependent explanations for the observed sex differences in RA presentation, including the influence of sex-hormones, discrepancies in the processing and reporting of pain, sex-specific applicability of disease activity measurements and bias in healthcare delivery^36,41^. These highlight the need to consider such factors when devising personalized management plans and treatment recommendations, and particularly when incorporating ML into practice due to its ability to exacerbate such differences.

*Investigating the extent to which sex bias is present in Machine Learning research in Rheumatoid Arthritis*

The objective of this systematic review is to investigate the extent to which sex bias is considered and mitigated in ML research related to RA. We focus on five distinct types of bias, specifically:

1. Training Data Bias, stemming from sex skew in the training dataset.
2. Test Data Bias, originating from sex skew in the test dataset.
3. Input Variable Bias, arising from input variables that may be biased in sex representation.
4. Output Variable Bias, associated with output variables that may be biased in sex representation,
5. Analysis Bias, which results from the omission of sex disaggregated analyses.

By examining these aspects, we seek to shed light on which of these potential biases are considered during ML model development in the domain of RA, how often they occur, and ultimately strive to improve responsible Artificial Intelligence (AI) and transparency in mitigating bias and ensuring equitable outcomes in healthcare.

## Methods

The reporting of this systematic review was guided by the standards of the Preferred Reporting Items for Systematic Review and Meta-Analysis (PRISMA) Statement. The protocol for this systematic review was pre-registered on PROSPERO with ID CRD42023431754.

### Search Strategy

The search strategy was defined to identify research studies using machine learning in the domain of rheumatoid arthritis from two electronic databases, PubMed and EMBASE. Search terms related to machine learning and rheumatoid arthritis were combined into the following search strategy: (machine learning OR ML OR artificial intelligence OR AI OR deep learning) AND (rheumatoid arthritis OR RA OR rheumato* arthrit* OR rheumato* joint inflamm*). Searches were restricted to articles in English, and published between January 2018 and May 2023 (when the searches were run) to capture the most recent five years of research in the rapidly developing field of machine learning research. References from relevant review papers were explored, and the search was rerun in November 2023 before finalising the review to identify additional papers.

### Study Selection

The inclusion and exclusion criteria (Table 1) were consistently and iteratively applied to filter search results. The review focused on supervised ML papers and biological sex (rather than sex and gender) reporting to support consistency in the application of the bias-checking matrix. Further, we included papers which applied ML in the context of prediction (due to their more immediate applicability in clinical practice) rather than on inferential statistical analysis (conclusions around associations or relationships between variables) and used this rule for papers implementing methods that can be used for both (e.g., linear and logistic regression).

**Table 1.**
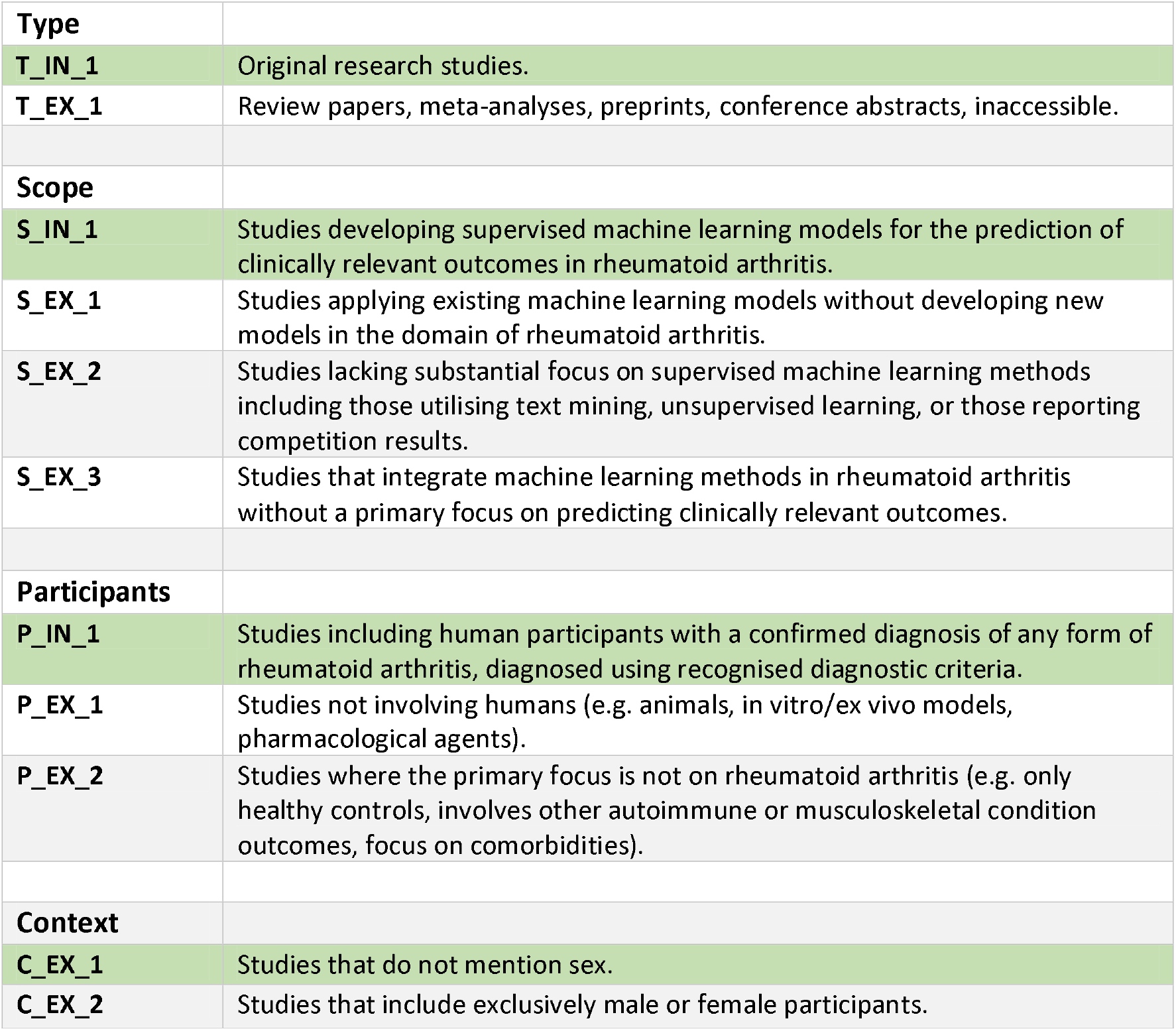
Inclusion and exclusion criteria for paper screening.

Search results were exported to Mendeley and duplicate entries removed. ST and CM conducted title and abstract screening to select relevant studies for further examination. AT and GQ independently screened the full-text version of the remaining papers for inclusion based on the eligibility criteria. Any disagreements between reviewers were resolved through discussion with SG. The PRISMA flow diagram (Figure 1) provides a visual representation of the study selection process and the reasons for exclusion at each stage. Forty-five papers were included from the initial search and screening, with a further two identified from review papers, and a further five identified when the searches were rerun.

**Figure 1.**
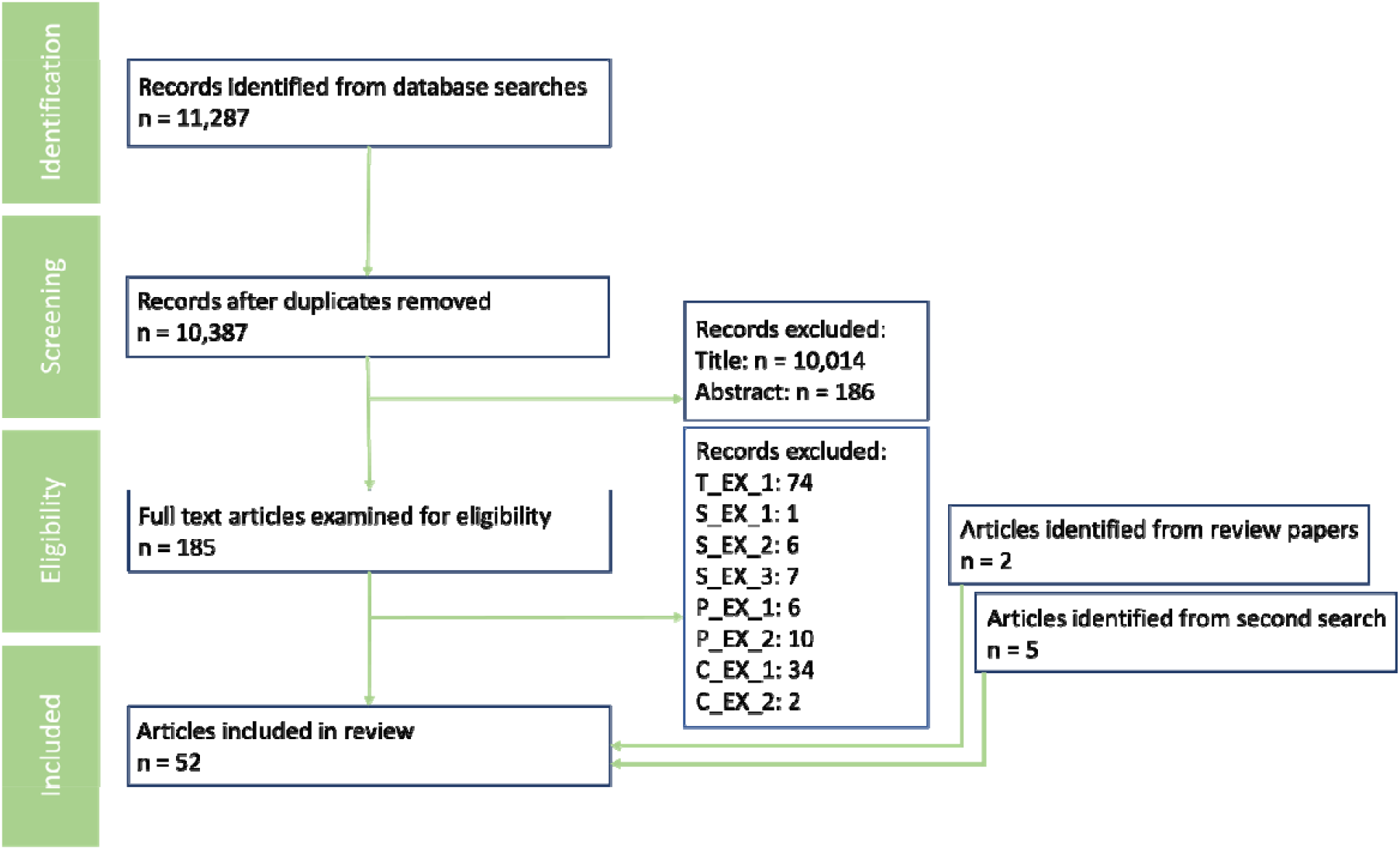
PRISMA flow diagram of study selection and screening process. See Table 1 for the exclusion code definitions.

AT, ST, CM and GQ completed the data extraction and quality assessment from each of the full-text papers with AT checking scores to ensure consistent interpretation of the criteria. Any conflicts in assessment were resolved through discussion with SG. All scores were recorded in an Excel spreadsheet (Table S1).

### Data Extraction

For each of the full-text papers, the study objectives, types of ML method used, and sex split of participants with RA were noted. Each paper was scored on whether the authors reported, attempted to mitigate or successfully mitigated the following types of bias: training data bias, test data bias, input variable bias, output variable bias, analysis bias, using a specifically designed scoring matrix (Table 2). Coherent with our criteria, included papers were required to provide participant’s sex. Thus, for training and test data bias, we expected authors to comment on the impact of sex imbalance on model results to score 1. However, for the input and output variable bias, and analysis bias, simply reporting how variables or analysis differed by sex was sufficient to score 1.

**Table 2.** Sex Bias Scoring Matrix.

| **Bias** | **Definition** |
| --- | --- |
| **Training Data** | sex skew in the dataset used to train the model |
| **Test Data** | sex skew in the dataset used to test the model |
| **Input Variable** | input variables that better represent a particular sex |
| **Output Variable** | output variables that better represent a particular sex |
| **Analysis** | disaggregation of model performance by sex |
| **Score** | **Definition** |
| **N/A** | not applicable |
| **0** | not mentioned |
| **1** | reported but not mitigated, unsuccessfully mitigated, or result of mitigation unclear |
| **2** | successfully mitigated |

Of note, it is now widely acknowledged that sex and gender are distinct but interconnected concepts with sex referring to the biological characteristics, primarily chromosomes, reproductive organs, and hormones, and gender referring to broader social, cultural, and psychological aspects an individual identifies with, which may correspond or not to the established ideas of male or female. In this paper, we refer to the sex of participants, as our variable of interest. However, in reviewing the literature, sex and gender terms are rarely distinguished or clarified, including whether these are cis-gender terms or inclusive of other gender individuals. In this paper, it is assumed that the sex characteristic reflects the biologic sex or that all participants are cis-gender due to the lack of clarity of all papers included in the final analysis.

### Quality Assessment

The quality assessment matrix (Table 3) was adapted and simplified from a previously published framework^42^. Papers were scored on their implementation of the following methodologies: pre-processing decisions, cross-validation, sample size, evaluation, hyperparameters, ensembling, reproducibility.

**Table 3.** Quality Assessment Matrix.

| Category | Score | Definition |
| --- | --- | --- |
| **Pre-processing decisions** | 0 | not reported |
|  | 1 | partially reported |
|  | 2 | fully reported and replicable |
| **Cross-Validation** | 0 | not done or used training data |
|  | 1 | used unseen sample from same dataset |
|  | 2 | used new sample |
| **Sample Size** | 0 | no calculation or mention of sample size |
|  | 1 | "big data" hinted at e.g. commented on size of sample |
|  | 2 | sample size calculation done |
| **Evaluation** | 0 | not done or accuracy metric only |
|  | 1 | accuracy metric plus error or calibration |
|  | 2 | all of accuracy, error and calibration |
| **Hyperparameters** | 0 | no information |
|  | 1 | performed manually, random search, or only report final values |
|  | 2 | optimised analytically or grid search |
| **Ensembling** | 0 | not applied |
|  | 1 | applied (including random forest, xgboost) but insufficiently justified |
|  | 2 | applied with clear justification |
| **Reproducibility** | 0 | none of data/code availability |
|  | 1 | some of data/code availability (statement or named dataset) |
|  | 2 | all of data/code availability |

### Data Synthesis

The objectives of each study were identified and used to classify papers accordingly. The ML algorithms used for clinical prediction in each paper were also extracted and classified into broad categories. Summary statistics, including mean and standard deviation, of the sex split of participants were calculated across 51 of the 52 reviewed papers (including one that additionally incorporated participants with undifferentiated arthritis with the primary focus of predicting their transition to RA^43^, and another that didn’t separate the sex split between the RA population with the healthy controls^44^). One paper was excluded from this analysis as it only reported the training dataset sex split^45^.

## Results

To investigate the extent to which sex bias is considered in machine learning for RA research, we conducted a search across two databases to identify studies from the most recent five years. The screening process resulted in the inclusion of 52 studies (Table S1) for qualitative synthesis and analysis.

### Characteristics of Included Papers

Data synthesis involved classifying the study objectives of papers. The resulting categories and their associated frequencies were: Predict Treatment Response (23), Score Disease Activity (11), Improve Diagnostic Accuracy (10), Assess Joint Damage (5), and Identify Patient Subgroups (3). The most frequent objective of included papers was improving treatment response predictions. This emphasis can be attributed to the intricate nature of clinical decision-making regarding therapeutic interventions and the significant potential of ML in refining precision medicine within this domain. This could also be a by-product of our inclusion criterion that mandated the reporting of participant sex. Such studies frequently employ symptom measurements derived directly from human participants, contrasting with those accessing blood or imaging results, which might explain this skewed distribution.

The ML techniques employed in each paper were also grouped into broad categories. The resulting categories and their associated frequencies were: Random Forest (30), Regression (25), Neural Networks (21), Support Vector Machine (17),Boosted Tree (15), K Nearest Neighbours (6), Naïve Bayes (4), Other (3, including Gaussian Process, Pathway Supported Models and Hidden Markov Models). Numerous papers implemented and compared the performance of multiple algorithms, with some techniques more popular for certain study objectives such as neural networks for scoring disease activity and assessing joint damage (Figure 2). In contrast, papers focused on improving diagnostic accuracy or identifying patient subgroups didn’t favour specific algorithms.

**Figure 2.**
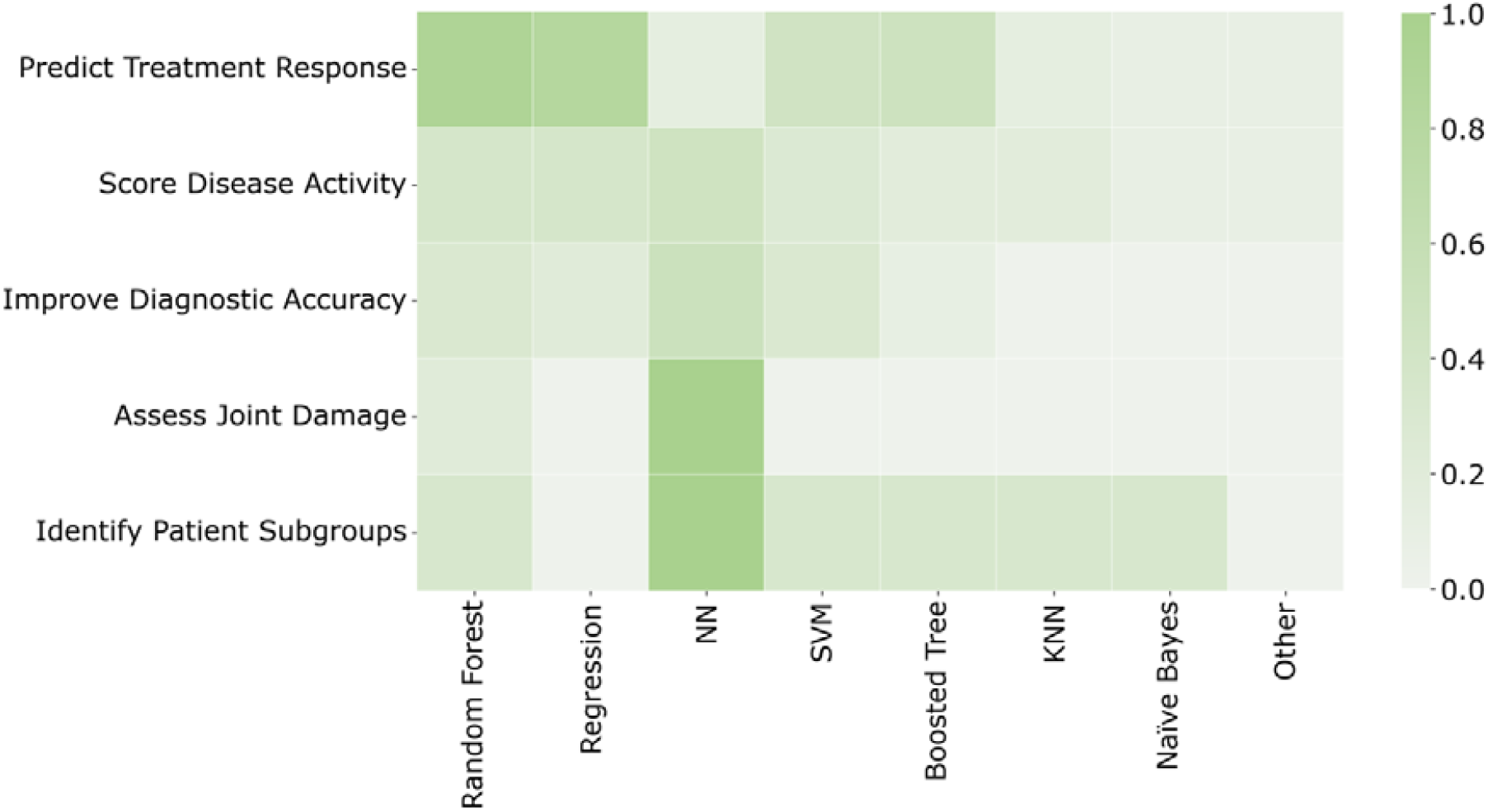
Proportion of papers with each study objective that used each type of ML model. NN: Neural Network, SVM: Support Vector Machine, KNN: K-Nearest Neighbours.

### Participant Sex Reporting and Participant Split

During full-text screening, 40 papers (six from the second search) that would otherwise have been in the scope of this review were excluded for not reporting the sex split of any RA participants (C_EX_1 in Table 1). A further two papers were excluded for including only female participants (C_EX_2 in Table 1). The mean (SD) of female participants in included papers was 74% (12%), consistent with the estimated global prevalence of rheumatoid arthritis at 70% female^20^. All papers had a female recruitment bias in their participants, with the exception of one study which only included 17% females^46^.

### Sex Bias Reporting and Mitigation

Forty-two papers scored 0 across all sex bias categories. The remaining ten papers scored a total of 1 across all categories. Details on scores for specific types of bias are given below.

### Training Data Bias & Test Data Bias

No papers included in the final review acknowledge that sex bias, in either the training or test data, can influence the sex bias of their models’ predictions.

### Input Variable Bias & Output Variable Bias

One paper statistically tested for sex differences in their model’s input variables, three bone biomarkers [osteopontin, stromelysin-1 (MMP3), and vascular endothelial growth factor-A (VEGF)] and reported no difference^47^. Six papers acknowledged potential output variable sex bias, five with the objective of evaluating treatment response and one improving disease activity scoring. Three papers mention sex-specific differences in baseline measures of erythrocyte sedimentation rate (ESR), often used as part of Disease Activity Scores (DAS), in their discussion section. Two of these suggest that women are less likely to achieve remission compared to men when using this measure^48,49^, and one suggests that lack of age and sex-specific cut-offs for ESR may bias results^50^. Three papers statistically tested for sex differences in their output variable: one reported that sex differed meaningfully between participants that achieved low disease activity (defined as clinical disease activity index - CDAI < 10), and those that didn’t^51^; the other two, however, found no difference between responders and non-responders to Adalimumab or Etanercept therapy at baseline for clinical parameters (including sex) in either of their two cohorts of data^52^, or to Leflunomide by sex^53^.

### Analysis Bias

Three papers assessed bias in model performance by sex disaggregating their results using different approaches. One study, aimed at improving diagnostic accuracy^46^, presented the percentage of male participants for each type of outcome: True Negative (57.9%), True Positive (81.8%), False Positive (75.0%) and False Negative (66.7%). The researchers used a dataset with 83.2% males in the RA group, and 59.4% males in the control group, which likely explains the higher representation of males in the positive outcomes (both true and false) compared to the negative ones. However, this information does not actually tell us whether the model is equally accurate for diagnosing males and females overall, so the implications of these representation skews is unclear. Another study^54^ presented the AUROC results for the detection of active synovitis in different age and sex groups. Males (n = 29) had an AUROC of 0.83 (95% CI, 0.67 to 0.99; p<0.01), whilst females (n = 117) had an AUROC 0.77 (95% CI, 0.68 to 0.85; p<0.01). Thus, despite having a strong female skew in the dataset, the authors described model performance as “similar” across sexes, despite not assessing this statistically. Finally, one study^55^, statistically tested sex differences in performance of classification and regression algorithms for predicting disease activity scores. Whilst classification was more accurate for males, and regression more accurate for women, these differences were not statistically significant. However, it demonstrates an interesting point, that even when trained on the same data, different models exhibit varying accuracy across sex-based subgroups.

### Quality Assessment

We conducted a quality assessment to evaluate the robustness of the data and the overall rigor of the research. A score of 0, 1 or 2 was given to each paper for each of seven quality metrics with equal weighting (Figure 3), thus giving a total quality score out of 14. Papers scoring 0-4 were deemed low quality, 5-9 medium and 10-14 considered high quality. Of the 47 papers, only one paper was scored as high quality, 38 were determined to be medium quality, with the remainder low quality. The most common score was a 1 which generally indicated that papers covered the method, but with insufficient detail or justification. The stand-out anomaly to this was sample size where most papers scored 0, in line with previous reports of this being an issue in ML research in RA^56^. There was no correlation between the quality of the papers and their consideration of sex bias (r = 0.06, p = 0.65), with the only high-quality paper scoring zero for sex bias consideration. ^56^

**Figure 3.**
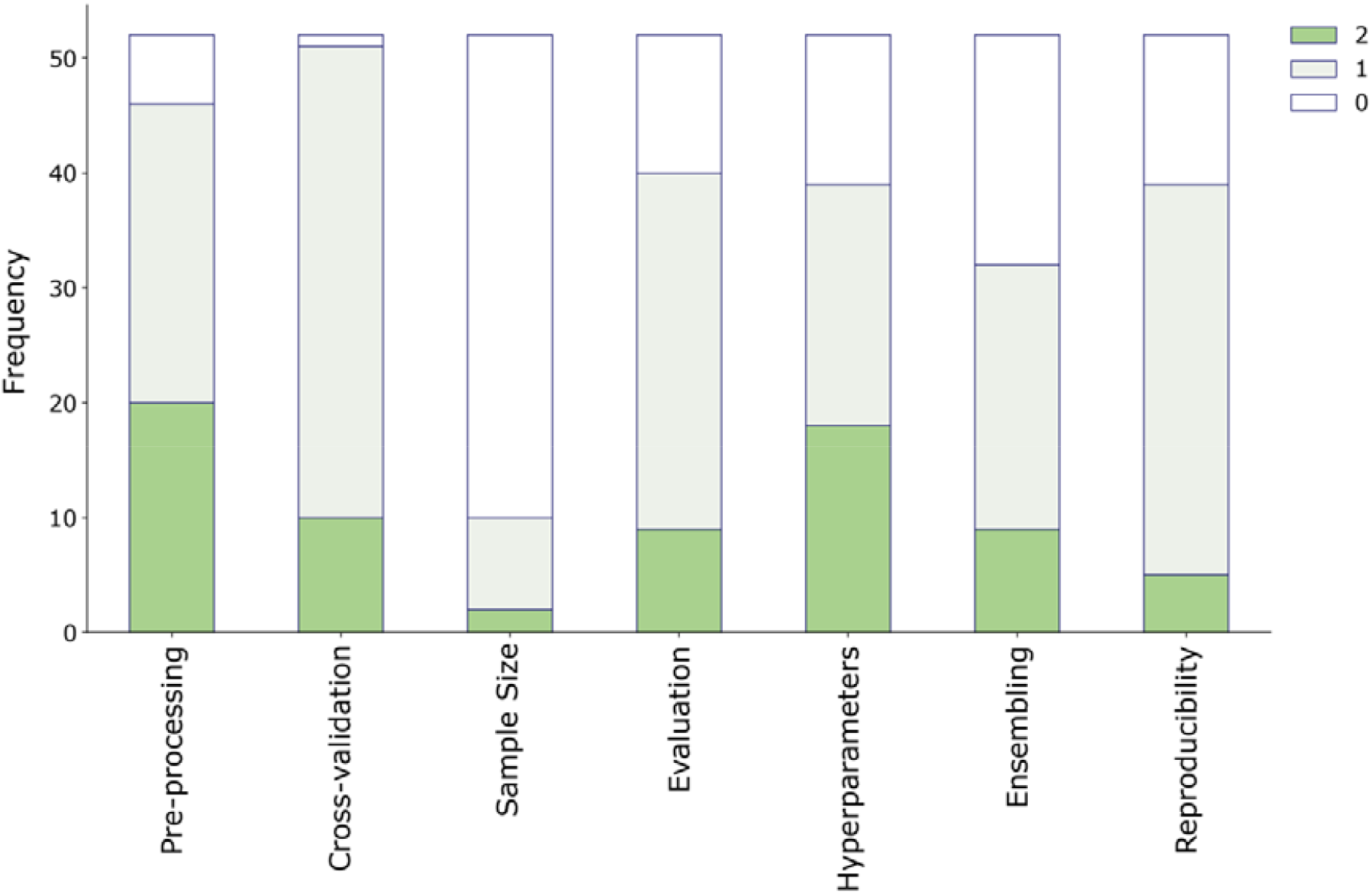
Number of papers scoring 0, 1 or 2 for each category in our quality assessment matrix.

## Discussion

We observed a substantial body of literature from EMBASE and PubMed pertaining to ML applied in RA. Fifty-two papers passed our screening criteria, to which we applied a specifically developed sex bias checking matrix. Forty-two research papers did not report or consider the potential for sex bias in any of our five categories: training data, test data, input variables, output variables or analysis. Only one reported potential sex bias in input variables, six in the output variables, whilst three disaggregated their model performance results by sex. No paper attempted to mitigate potential sex biases in their model development process. Our analysis demonstrates that sex bias is generally not considered in the development of ML models and raises important questions about how its clinical application might contribute to or mitigate sex bias in healthcare.

Analysis bias, which involves assessing model performance by sex, rather than using a composite metric, is arguably the most impactful of the five types of bias we assessed. It is essential for demonstrating the presence and direction of bias and allows us to understand the extent of impact, if any, of data skews or biased model variables, as well as the efficacy of mitigation methods. Only three papers of the 52 reviewed disaggregated model performance by sex, though the differences were generally not tested statistically, and with minimal explanation attached to their findings.

A major finding is that no paper commented on the potential impacts of sex skews in data, despite a mean sex split of 74% female participants across included papers. This information was readily available to the authors as papers had to provide the sex split of RA patients to be included in our review. One likely explanation for the dearth of data bias reporting is that most researchers considered their female-skewed data to be representative of the general RA population, thereby mirroring FDA guidance for clinical trials (particularly as ‘predicting treatment response’ was the most common objective)^57^. Regardless, it is known that sex imbalance in data generally makes results less applicable to the minority groups, a discrepancy which can be further exacerbated in ML research^19,58^. Furthermore, it is noteworthy that one of the papers analysed contained a dataset in which only 17% of the participants were female^46^, in sharp contrast with the reported sex prevalence. Additionally, three other papers relied on datasets with an overwhelming female representation (over 85%)^59–61^. The absence of acknowledgment of the impact of skewed sex distributions on the representativeness of algorithmic predictions within these papers, despite easily implementable data balancing mitigation methods^19,58,62,63^, suggests an element of limited awareness or under prioritisation of fair and equitable models.

Whilst one paper assessed and reports no sex difference in the model’s input variables, six papers recognised potential sex bias in their output variables, specifically regarding a skew to higher scores in females on the disease activity measures. This observation has been previously reported in the RA literature and is believed to be rooted in the self-report nature of the composite measure, rather than the physical or physiological aspects it encompasses^36^. The inclination for women to report higher pain scores has been extensively documented, yet the underlying causes remain unclear. It’s possible that this divergence in self-reported scores arises from a combination of poorly understood physiological and psychological factors^64,65^. The challenge lies in distinguishing between sex bias in measurement and genuine sex-based differences in disease activity within the general population. Addressing this complexity may involve using carefully constructed and validated measurement tools and implementing modelling strategies to ensure predictive accuracy remains equivalent between the sexes.

We acknowledge several limitations of the research methods and results. Firstly, the nature of a systematic review introduces the potential for selection bias. Publication bias means that we may not have captured all available studies in the field. The search strategy, while comprehensive, may miss studies due to variations in terminologies and databases. Additionally, the review was limited to papers published in the English language, which may have excluded relevant non-English studies. Furthermore, the diversity in study methods, measures, and study populations across the papers we reviewed introduced significant heterogeneity. This made it challenging to apply our inclusion/exclusion criteria consistently across the entire dataset. Similarly, the types of bias assessed were not complete^66^, but enabled consistency in our review. Another key limitation pertains to the limited explanation of sex and gender features within almost all papers included in our analysis, making it difficult to know exactly how this information was collected and what it represents, and may have led to conflation of these two distinct concepts. Finally, it is essential to acknowledge that this review focused primarily on one type of bias, namely sex bias. However, the development of fair and equitable algorithms in healthcare and machine learning involves multiple facets, including those related to race, socioeconomic status, and more. This should be considered in the development of ML legislation, such as the European Union’s Artificial Intelligence act^67^, to encourage disaggregating analyses by specific subgroups. Thus, while the study aimed to shed light on the specific issue of sex bias in the context of RA, a comprehensive assessment of bias in healthcare algorithms demands a broader perspective.

This systematic review invites further exploration into whether current research approaches are inadvertently perpetuating sex bias, through their clinical impact. Our findings underscore the necessity of considering broader objectives and diversifying the focus of machine learning applications in healthcare, to also promoting inclusive and equitable healthcare solutions.

## Data Availability

All data produced in the present work are contained in the manuscript.

## Acknowledgements

We extend our sincere gratitude to Dr Billy Franks for their insightful review of this manuscript. Funding for research wholly derived from HALEON standing R&D. There were no additional sponsors or funding bodies.

## Data Availability

All data extracted from papers and their corresponding references, are comprehensively documented and presented in Supplementary Table 1. Researchers interested in accessing the raw data for verification or further analysis can refer to Supplementary Table 1 for a detailed compilation.

