## Supplemental Table 1 for "Investigating Sex Bias in Machine Learning Research: A Systematic Review in Rheumatoid Arthritis"

| No. | First Author | Title | Year | Study Objective | ML Models | Overall Sex Split % Female RA | Training Data Female RA % | Test Data Female RA % | Training data Bias | Test Data Bias | Input Variable Bias | Output Variable Bias | Analysis Bias | Total (X/10) | Pre-processing | Cross-validation | Sample Size | Evaluation | Hyperparameters | Ensembling | Reproducibility | Total (X/14) | Reference |
|---|---|---|---|---|---|---|---|---|---|---|---|---|---|---|---|---|---|---|---|---|---|---|---|
| 1 | Bai | Improved diagnosis of rheumatoid arthritis using an artificial neural network | 2022 | Improve Diagnostic Accuracy | NN | 17 | NA | NA | 0 | 0 | 0 | 0 | 1 | 1 | 1 | 1 | 0 | 1 | 2 | 1 | 0 | 6 | Bai, L., Zhang, Y., Wang, P., Zhu, X., Xiong, J. W., & Cui, L. (2022). Improved diagnosis of rheumatoid arthritis using an artificial neural network. Scientific Reports, 12(1). https://doi.org/10.1038/S41598-022-13750-9 |
| 2 | Bouget | Machine learning predicts response to TNF inhibitors in rheumatoid arthritis: results on the ESPOIR and ABIRISK cohorts | 2022 | Predict Treatment Response | Regression, Random Forest, Boosted Tree | 71 | 71 | 72 | 0 | 0 | 0 | 0 | 0 | 0 | 1 | 2 | 0 | 2 | 0 | 2 | 1 | 8 | Bouget, V., Duquesne, J., Hassler, S., Cournède, P.-H., Fautrel, B., Guillemin, F., Pallardy, M., Broët, P., Mariette, X., & Bitoun, S. (2022). Machine learning predicts response to TNF inhibitors in rheumatoid arthritis: results on the ESPOIR and ABIRISK cohorts. RMD Open, 8(2). https://doi.org/10.1136/rmdopen-2022-002442 |
| 3 | Chin | EDram: Effective early disease risk assessment with matrix factorization on a large-scale medical database: A case study on rheumatoid arthritis | 2018 | Improve Diagnostic Accuracy | SVM | 76 | NA | NA | 0 | 0 | 0 | 0 | 0 | 0 | 2 | 1 | 0 | 1 | 2 | 0 | 1 | 7 | Chin, C.-Y., Hsieh, S.-Y., & Tseng, V. S. (2018). EDram: Effective early disease risk assessment with matrix factorization on a large-scale medical database: A case study on rheumatoid arthritis. PLoS ONE, 13(11). https://doi.org/10.1371/journal.pone.0207579 |
| 4 | Chocholova | Glycomics meets artificial intelligence – Potential of glycan analysis for identification of seropositive and seronegative rheumatoid arthritis patients revealed | 2018 | Identify Patient Subgroups | NN | 83 | NA | NA | 0 | 0 | 0 | 0 | 0 | 0 | 2 | 1 | 0 | 0 | 1 | 0 | 0 | 4 | Chocholova, E., Bertok, T., Jane, E., Lorencova, L., Holazova, A., Belicka, L., Belicky, S., Mislovicova, D., Vikartovska, A., Imrich, R., Kasak, P., & Tkac, J. (2018). Glycomics meets artificial intelligence – Potential of glycan analysis for identification of seropositive and seronegative rheumatoid arthritis patients revealed. Clinica Chimica Acta, 481, 49–55. https://doi.org/10.1016/j.cca.2018.02.031 |
| 5 | Christensen | Applying cascaded convolutional neural network design further enhances automatic scoring of arthritis disease activity on ultrasound images from rheumatoid arthritis patients | 2020 | Score Disease Activity | NN | 68 | NA | NA | 0 | 0 | 0 | 0 | 0 | 0 | 2 | 1 | 0 | 0 | 0 | 0 | 0 | 3 | Christensen, A. B. H., Just, S. A., Andersen, J. K. H., & Savarimuthu, T. R. (2020). Applying cascaded convolutional neural network design further enhances automatic scoring of arthritis disease activity on ultrasound images from rheumatoid arthritis patients. Annals of the Rheumatic Diseases, 79(9), 1189–1193. https://doi.org/10.1136/annrheumdis-2019-216636 |
| 6 | Cuppen | Proteomics to predict the response to tumour necrosis factor-α inhibitors in rheumatoid arthritis using a supervised cluster-analysis based protein score | 2018 | Predict Treatment Response | Regression | 74 | 72 | 75 | 0 | 0 | 0 | 0 | 0 | 0 | 2 | 1 | 0 | 2 | 0 | 0 | 1 | 6 | Cuppen, B. V. J., Fritsch-Stork, R. D. E., Eekhout, I., de Jager, W., Marijnissen, A. C., Bijlsma, J. W. J., Custers, M., van Laar, J. M., Lafeber, F. P. J. G., & Welsing, P. M. J. (2018). Proteomics to predict the response to tumour necrosis factor-α inhibitors in rheumatoid arthritis using a supervised cluster-analysis based protein score. Scandinavian Journal of Rheumatology, 47(1), 12–21. https://doi.org/10.1080/03009742.2017.1309061 |
| 7 | Curtis | Machine Learning Applied to Patient-Reported Outcomes to Classify Physician-Derived Measures of Rheumatoid Arthritis Disease Activity | 2022 | Score Disease Activity | Random Forest, Boosted Tree, SVM, Regression | 83 | NA | NA | 0 | 0 | 0 | 1 | 0 | 1 | 1 | 1 | 0 | 1 | 1 | 1 | 1 | 6 | Curtis, J. R., Su, Y., Black, S., Xu, S., Langholff, W., Bingham, C. O., Kafka, S., & Xie, F. (2022). Machine Learning Applied to Patient-Reported Outcomes to Classify Physician-Derived Measures of Rheumatoid Arthritis Disease Activity. ACR Open Rheumatology, 4(12), 995–1003. https://doi.org/10.1002/acr2.11499 |
| 8 | de la Calle-Fabregat | Prediction of the Progression of Undifferentiated Arthritis to Rheumatoid Arthritis Using DNA Methylation Profiling | 2021 | Improve Diagnostic Accuracy | Regression, Random Forest, SVM | 57 | 58 | 56 | 0 | 0 | 0 | 0 | 0 | 0 | 2 | 2 | 0 | 1 | 0 | 1 | 1 | 7 | de la Calle-Fabregat, C., Niemantsverdriet, E., Cañete, J. D., Li, T., van der Helm-van Mil, A. H. M., Rodríguez-Ubreva, J., & Ballestar, E. (2021). Prediction of the Progression of Undifferentiated Arthritis to Rheumatoid Arthritis Using DNA Methylation Profiling. Arthritis and Rheumatology, 73(12), 2229–2239. https://doi.org/10.1002/art.41885 |
| 9 | Duong | Clinical predictors of response to methotrexate in patients with rheumatoid arthritis: a machine learning approach using clinical trial data | 2022 | Predict Treatment Response | Regression, Random Forest | 80 | 78 | 81 | 0 | 0 | 0 | 1 | 0 | 1 | 1 | 2 | 1 | 2 | 0 | 2 | 1 | 9 | Duong, S. Q., Crowson, C. S., Athreya, A., Atkinson, E. J., Davis, J. M., Warrington, K. J., Matteson, E. L., Weinshilboum, R., Wang, L., & Myasoedova, E. (2022). Clinical predictors of response to methotrexate in patients with rheumatoid arthritis: a machine learning approach using clinical trial data. Arthritis Research and Therapy, 24(1). https://doi.org/10.1186/s13075-022-02851-5 |
| 10 | Duquesne | Machine learning identifies a profile of inadequate responder to methotrexate in rheumatoid arthritis | 2022 | Predict Treatment Response | Regression, Random Forest, Boosted Tree | 71 | 70 | 72 | 0 | 0 | 0 | 0 | 0 | 0 | 2 | 2 | 1 | 2 | 0 | 1 | 1 | 9 | Duquesne, J., Bouget, V., Cournède, P. H., Fautrel, B., Guillemin, F., de Jong, P. H. P., Heutz, J. W., Verstappen, M., van der Helm-van Mil, A. H. M., Mariette, X., & Bitoun, S. (2022). Machine learning identifies a profile of inadequate responder to methotrexate in rheumatoid arthritis. Rheumatology (Oxford, England). https://doi.org/10.1093/rheumatology/keac645 |
| 11 | Feldman | Supplementing Claims Data with Electronic Medical Records to Improve Estimation and Classification of Rheumatoid Arthritis Disease Activity: A Machine Learning Approach | 2019 | Score Disease Activity | Regression | 80 | NA | NA | 0 | 0 | 0 | 0 | 0 | 0 | 1 | 1 | 0 | 1 | 2 | 0 | 1 | 6 | Feldman, C. H., Yoshida, K., Xu, C., Frits, M. L., Shadick, N. A., Weinblatt, M. E., Connolly, S. E., Alemao, E., & Solomon, D. H. (2019). Supplementing Claims Data with Electronic Medical Records to Improve Estimation and Classification of Rheumatoid Arthritis Disease Activity: A Machine Learning Approach. ACR Open Rheumatology, 1(9), 552–559. https://doi.org/10.1002/acr2.11068 |
| 12 | Folle | Deep Learning-Based Classification of Inflammatory Arthritis by Identification of Joint Shape Patterns—How Neural Networks Can Tell Us Where to "Deep Dive" Clinically | 2022 | Improve Diagnostic Accuracy | NN | 69 | NA | NA | 0 | 0 | 0 | 0 | 0 | 0 | 2 | 1 | 0 | 1 | 0 | 0 | 1 | 5 | Folle, L., Bayat, S., Kleyer, A., Fagni, F., Kapsner, L. A., Schlereth, M., Meinderink, T., Breininger, K., Tascilar, K., Krönke, G., Uder, M., Sticherling, M., Bickelhaupt, S., Schett, G., Maier, A., Roemer, F., & Simon, D. (2022). Advanced neural networks for classification of MRI in psoriatic arthritis, seronegative, and seropositive rheumatoid arthritis. Rheumatology (Oxford, England), 61(12), 4945–4951. https://doi.org/10.1093/RHEUMATOLOGY/KEAC197 |

| | | | | | | | | | | | | | | | | | | | | | | | |
|---|---|---|---|---|---|---|---|---|---|---|---|---|---|---|---|---|---|---|---|---|---|---|---|
| 13 | Folle | Advanced neural networks for classification of MRI in psoriatic arthritis, seronegative, and seropositive rheumatoid arthritis | 2022 | Identify Patient Subgroups | NN | 67 | NA | NA | 0 | 0 | 0 | 0 | 0 | 0 | 0 | 1 | 0 | 1 | 1 | 0 | 2 | 5 | Folle, L., Simon, D., Tascilar, K., Krönke, G., Liphardt, A.-M., Maier, A., Schett, G., & Kleyer, A. (2022). Deep Learning-Based Classification of Inflammatory Arthritis by Identification of Joint Shape Patterns—How Neural Networks Can Tell Us Where to "Deep Dive" Clinically. Frontiers in Medicine, 9. https://doi.org/10.3389/fmed.2022.850552 |
| 14 | Fukae | Convolutional neural network for classification of two-dimensional array images generated from clinical information may support diagnosis of rheumatoid arthritis | 2020 | Improve Diagnostic Accuracy | NN | 60 | 60 | 60 | 0 | 0 | 0 | 0 | 0 | 0 | 1 | 1 | 0 | 0 | 1 | 0 | 0 | 3 | Fukae, J., Isobe, M., Hattori, T., Fujieda, Y., Kono, M., Abe, N., Kitano, A., Narita, A., Henmi, M., Sakamoto, F., Aoki, Y., Ito, T., Mitsuzaki, A., Matsuhashi, M., Shimizu, M., Tanimura, K., Sutherland, K., Kamishima, T., Atsumi, T., & Koike, T. (2020). Convolutional neural network for classification of two-dimensional array images generated from clinical information may support diagnosis of rheumatoid arthritis. Scientific Reports, 10(1), 5648. https://doi.org/10.1038/s41598-020-62634-3 |
| 15 | Geng | Prediction of diagnosis results of rheumatoid arthritis patients based on autoantibodies and cost-sensitive neural network | 2022 | Improve Diagnostic Accuracy | NN | 78 | NA | NA | 0 | 0 | 0 | 0 | 0 | 0 | 1 | 1 | 0 | 0 | 1 | 0 | 1 | 4 | Geng, L., Qu, W., Wang, S., Chen, J., Xu, Y., Kong, W., Xu, X., Feng, X., Zhao, C., Liang, J., Zhang, H., & Sun, L. (2022). Prediction of diagnosis results of rheumatoid arthritis patients based on autoantibodies and cost-sensitive neural network. Clinical Rheumatology, 41(8), 2329–2339. https://doi.org/10.1007/s10067-022-06109-y |
| 16 | Gossec | Detection of Flares by Decrease in Physical Activity, Collected Using Wearable Activity Trackers in Rheumatoid Arthritis or Axial Spondyloarthritis: An Application of Machine Learning Analyses in Rheumatology | 2019 | Score Disease Activity | Naïve Bayes | 83 | NA | NA | 0 | 0 | 0 | 0 | 0 | 0 | 2 | 1 | 0 | 1 | 0 | 0 | 1 | 5 | Gossec, L., Guyard, F., Leroy, D., Lafargue, T., Seiler, M., Jacquemin, C., Molto, A., Sellam, J., Foltz, V., Gandjbakhch, F., Hudry, C., Mitrovic, S., Fautrel, B., & Servy, H. (2019). Detection of Flares by Decrease in Physical Activity, Collected Using Wearable Activity Trackers in Rheumatoid Arthritis or Axial Spondyloarthritis: An Application of Machine Learning Analyses in Rheumatology. Arthritis Care and Research, 71(10), 1336–1343. https://doi.org/10.1002/acr.23768 |
| 17 | Gosselt | Complex machine-learning algorithms and multivariable logistic regression on par in the prediction of insufficient clinical response to methotrexate in rheumatoid arthritis | 2021 | Predict Treatment Response | Regression, Random Forest, Boosted Tree | 68 | NA | NA | 0 | 0 | 0 | 0 | 0 | 0 | 1 | 1 | 2 | 2 | 1 | 1 | 1 | 9 | Gosselt, H. R., Verhoeven, M. M. A., Bulatović-ćalasan, M., Welsing, P. M., de Rotte, M. C. F. J., Hazes, J. M. W., Lafeber, F. P. J. G., Hoogendoorn, M., & de Jonge, R. (2021). Complex machine-learning algorithms and multivariable logistic regression on par in the prediction of insufficient clinical response to methotrexate in rheumatoid arthritis. Journal of Personalized Medicine, 11(1), 1–12. https://doi.org/10.3390/jpm11010044 |
| 18 | Guan | Machine Learning to Predict Anti–Tumor Necrosis Factor Drug Responses of Rheumatoid Arthritis Patients by Integrating Clinical and Genetic Markers | 2019 | Predict Treatment Response | Random Forest, Boosted Tree, Regression, SVM, Other | 76 | 75 | 78 | 0 | 0 | 0 | 0 | 0 | 0 | 0 | 2 | 0 | 1 | 2 | 1 | 1 | 7 | Guan, Y., Zhang, H., Quang, D., Wang, Z., Parker, S. C. J., Pappas, D. A., Kremer, J. M., & Zhu, F. (2019). Machine Learning to Predict Anti–Tumor Necrosis Factor Drug Responses of Rheumatoid Arthritis Patients by Integrating Clinical and Genetic Markers. Arthritis and Rheumatology, 71(12), 1987–1996. https://doi.org/10.1002/art.41056 |
| 19 | Hirano | Development and validation of a deep-learning model for scoring of radiographic finger joint destruction in rheumatoid arthritis | 2019 | Assess Joint Damage | NN | 83 | 83 | 87 | 0 | 0 | 0 | 0 | 0 | 0 | 1 | 1 | 0 | 0 | 1 | 0 | 0 | 3 | Hirano, T., Nishide, M., Nonaka, N., Seita, J., Ebina, K., Sakurada, K., & Kumanogoh, A. (2019). Development and validation of a deep-learning model for scoring of radiographic finger joint destruction in rheumatoid arthritis. Rheumatology Advances in Practice, 3(2). https://doi.org/10.1093/rap/rkz047 |
| 20 | Honda | Development of a scoring model for the Sharp/van der Heijde score using convolutional neural networks and its clinical application | 2022 | Assess Joint Damage | NN, Random Forest | 89 | NA | NA | 0 | 0 | 0 | 0 | 0 | 0 | 2 | 1 | 1 | 1 | 1 | 1 | 1 | 8 | Honda, S., Yano, K., Tanaka, E., Ikari, K., & Harigai, M. (2022). Development of a scoring model for the Sharp/van der Heijde score using convolutional neural networks and its clinical application. Rheumatology (Oxford, England). https://doi.org/10.1093/rheumatology/keac586 |
| 21 | Johansson | Predicting response to tocilizumab monotherapy in rheumatoid arthritis: A real-world data analysis using machine learning | 2021 | Predict Treatment Response | Regression, Random Forest | 76 | NA | NA | 0 | 0 | 0 | 0 | 0 | 0 | 1 | 1 | 0 | 2 | 0 | 2 | 0 | 6 | Johansson, F. D., Collins, J. E., Yau, V., Guan, H., Kim, S. C., Losina, E., Sontag, D., Stratton, J., Trinh, H., Greenberg, J., & Solomon, D. H. (2021). Predicting response to tocilizumab monotherapy in rheumatoid arthritis: A real-world data analysis using machine learning. Journal of Rheumatology, 48(9), 1364–1370. https://doi.org/10.3899/jrheum.201626 |
| 22 | Kalweit | Personalized prediction of disease activity in patients with rheumatoid arthritis using an adaptive deep neural network | 2021 | Score Disease Activity | NN, Random Forest, Regression, SVM | 74 | NA | NA | 0 | 0 | 0 | 0 | 1 | 1 | 1 | 1 | 0 | 1 | 1 | 1 | 1 | 6 | Kalweit, M., Walker, U. A., Finckh, A., Müller, R., Kalweit, G., Scherer, A., Boedecker, J., & Hügle, T. (2021). Personalized prediction of disease activity in patients with rheumatoid arthritis using an adaptive deep neural network. PLoS ONE, 16(6 June). https://doi.org/10.1371/journal.pone.0252289 |
| 23 | Kim | Effects of RETN polymorphisms on treatment response in rheumatoid arthritis patients receiving TNF-α inhibitors and utilization of machine-learning algorithms | 2022 | Predict Treatment Response | Regression, Random Forest, SVM | 81 | NA | NA | 0 | 0 | 0 | 0 | 0 | 0 | 0 | 1 | 0 | 1 | 2 | 1 | 1 | 6 | Kim, W., Jin Oh, S., Thi Trinh, N., Yeon Gil, J., Ah Choi, I., Hyoun Kim, J., Hee Kim, J., Jung, J.-Y., Kim, J., Kim, H.-A., & Eun Lee, K. (2022). Effects of RETN polymorphisms on treatment response in rheumatoid arthritis patients receiving TNF-α inhibitors and utilization of machine-learning algorithms. International Immunopharmacology, 111. https://doi.org/10.1016/j.intimp.2022.109094 |
| 24 | Kim | Association of TLR 9 gene polymorphisms with remission in patients with rheumatoid arthritis receiving TNF-α inhibitors and development of machine learning models | 2021 | Predict Treatment Response | Random Forest, SVM, Regression | 81 | NA | NA | 0 | 0 | 0 | 0 | 0 | 0 | 2 | 1 | 0 | 1 | 2 | 1 | 1 | 8 | Kim, W., Kim, T. H., Oh, S. J., Kim, H. J., Kim, J. H., Kim, H.-A., Jung, J.-Y., Choi, I. A., & Lee, K. E. (2021). Association of TLR 9 gene polymorphisms with remission in patients with rheumatoid arthritis receiving TNF-α inhibitors and development of machine learning models. Scientific Reports, 11(1), 20169. https://doi.org/10.1038/s41598-021-99625-x |

| | | | | | | | | | | | | | | | | | | | | | |
|---|---|---|---|---|---|---|---|---|---|---|---|---|---|---|---|---|---|---|---|---|---|
| 25 | Kim | Association between SYVN1 and SEL1 genetic polymorphisms and remission in rheumatoid arthritis patients treated with TNF-α inhibitors: a machine learning approach | 2023 | Predict Treatment Response | Regression, Random Forest, SVM | 81 | NA | NA | 0 | 0 | 0 | 0 | 0 | 0 | 2 | 1 | 0 | 1 | 2 | 1 | 1 | 8 | Kim, W., Yeon, H. R., Kim, J. H., Kim, J. H., Kim, J. H., Kim, H.-A., Jung, J.-Y., Kim, J., Choi, I. A., & Lee, K. E. (2023). Association between SYVN1 and SEL1 genetic polymorphisms and remission in rheumatoid arthritis patients treated with TNF-α inhibitors: a machine learning approach. Immunologic Research. https://doi.org/10.1007/s12026-023-09382-4 |
| 26 | Koo | Machine learning model for identifying important clinical features for predicting remission in patients with rheumatoid arthritis treated with biologics | 2021 | Predict Treatment Response | Regression, SVM, Random Forest, Boosted Tree | 83 | NA | NA | 0 | 0 | 0 | 0 | 0 | 0 | 1 | 1 | 0 | 0 | 2 | 1 | 1 | 6 | Koo, B. S., Eun, S., Shin, K., Yoon, H., Hong, C., Kim, D.-H., Hong, S., Kim, Y.-G., Lee, C.-K., Yoo, B., & Oh, J. S. (2021). Machine learning model for identifying important clinical features for predicting remission in patients with rheumatoid arthritis treated with biologics. Arthritis Research and Therapy, 23(1). https://doi.org/10.1186/s13075-021-02567-y |
| 27 | Lee | Machine learning-based prediction model for responses of bDMARDs in patients with rheumatoid arthritis and ankylosing spondylitis | 2021 | Predict Treatment Response | Random Forest, Boosted Tree, SVM, NN | 84 | NA | NA | 0 | 0 | 0 | 0 | 0 | 0 | 0 | 2 | 0 | 1 | 2 | 2 | 2 | 9 | Lee, S., Kang, S., Eun, Y., Won, H.-H., Kim, H., Lee, J., Koh, E.-M., & Cha, H.-S. (2021). Machine learning-based prediction model for responses of bDMARDs in patients with rheumatoid arthritis and ankylosing spondylitis. Arthritis Research and Therapy, 23(1). https://doi.org/10.1186/s13075-021-02635-3 |
| 28 | Lim | Functional coding haplotypes and machine-learning feature elimination identifies predictors of Methotrexate Response in Rheumatoid Arthritis patients | 2022 | Predict Treatment Response | NN, SVM, Regression, Random Forest, Boosted Tree | 82 | NA | NA | 0 | 0 | 0 | 0 | 0 | 0 | 1 | 1 | 0 | 1 | 1 | 1 | 1 | 6 | Lim, A. J. W., Lim, L. J., Ooi, B. N. S., Koh, E. T., Tan, J. W. L., Chong, S. S., Khor, C. C., Tucker-Kellogg, L., Leong, K. P., & Lee, C. G. (2022). Functional coding haplotypes and machine-learning feature elimination identifies predictors of Methotrexate Response in Rheumatoid Arthritis patients. EBioMedicine, 75. https://doi.org/10.1016/j.ebiom.2021.103800 |
| 29 | Lim | Machine learning using genetic and clinical data identifies a signature that robustly predicts methotrexate response in rheumatoid arthritis | 2022 | Predict Treatment Response | SVM, NN, Random Forest, Boosted Tree, Regression | 82 | 82 | 84 | 0 | 0 | 0 | 0 | 0 | 0 | 2 | 1 | 0 | 1 | 1 | 1 | 1 | 7 | Lim, L. J., Lim, A. J. W., Ooi, B. N. S., Tan, J. W. L., Koh, E. T., Chong, S. S., Khor, C. C., Tucker-Kellogg, L., Lee, C. G., & Leong, K. P. (2022). Machine learning using genetic and clinical data identifies a signature that robustly predicts methotrexate response in rheumatoid arthritis. Rheumatology (United Kingdom), 61(10), 4175–4186. https://doi.org/10.1093/rheumatology/keac032 |
| 30 | Lötsch | Machine-learning-based knowledge discovery in rheumatoid arthritis-related registry data to identify predictors of persistent pain | 2020 | Identify Patient Subgroups | Random Forest, KNN, SVM, Naïve Bayes, Boosted Tree, NN | 73 | NA | NA | 0 | 0 | 0 | 0 | 0 | 0 | 2 | 1 | 1 | 0 | 2 | 0 | 1 | 7 | Lötsch, J., Alfredsson, L., & Lampa, J. (2020). Machine-learning-based knowledge discovery in rheumatoid arthritis-related registry data to identify predictors of persistent pain. Pain, 161(1), 114–126. https://doi.org/10.1097/j.pain.0000000000001693 |
| 31 | Maciejewski | Prediction of response of methotrexate in patients with rheumatoid arthritis using serum lipidomics | 2021 | Predict Treatment Response | SVM, Random Forest, Regression | 80 | NA | NA | 0 | 0 | 0 | 0 | 0 | 0 | 1 | 1 | 0 | 1 | 2 | 1 | 1 | 7 | Maciejewski, M., Sands, C., Nair, N., Ling, S., Verstappen, S., Hyrich, K., Barton, A., Ziemek, D., Lewis, M. R., & Plant, D. (2021). Prediction of response of methotrexate in patients with rheumatoid arthritis using serum lipidomics. Scientific Reports, 11(1), 7266. https://doi.org/10.1038/s41598-021-86729-7 |
| 32 | Matsuo | Machine learning-based prediction of relapse in rheumatoid arthritis patients using data on ultrasound examination and blood test | 2022 | Predict Treatment Response | Regression, Random Forest, Boosted Tree | 82 | NA | NA | 0 | 0 | 0 | 0 | 0 | 0 | 1 | 1 | 0 | 1 | 2 | 2 | 0 | 7 | Matsuo, H., Kamada, M., Imamura, A., Shimizu, M., Inagaki, M., Tsuji, Y., Hashimoto, M., Tanaka, M., Ito, H., & Fujii, Y. (2022). Machine learning-based prediction of relapse in rheumatoid arthritis patients using data on ultrasound examination and blood test. Scientific Reports, 12(1), 7224. https://doi.org/10.1038/s41598-022-11361-y |
| 33 | Mehta | Machine learning identification of thresholds to discriminate osteoarthritis and rheumatoid arthritis synovial inflammation | 2023 | Improve Diagnostic Accuracy | Random Forest | 83 | NA | NA | 0 | 0 | 0 | 0 | 0 | 0 | 1 | 1 | 0 | 1 | 2 | 1 | 1 | 7 | Mehta, B., Goodman, S., DiCarlo, E., Jannat-Khah, D., Gibbons, J. A. B., Otero, M., Donlin, L., Pannellini, T., Robinson, W. H., Sculco, P., Figgie, M., Rodriguez, J., Kirschmann, J. M., Thompson, J., Slater, D., Frezza, D., Xu, Z., Wang, F., & Orange, D. E. (2023). Machine learning identification of thresholds to discriminate osteoarthritis and rheumatoid arthritis synovial inflammation. Arthritis Research and Therapy, 25(1). https://doi.org/10.1186/s13075-023-03008-8 |
| 34 | Morales-Ivorra | Assessment of inflammation in patients with rheumatoid arthritis using thermography and machine learning: A fast and automated technique | 2022 | Score Disease Activity | KNN | 77 | 75 | 80 | 0 | 0 | 0 | 0 | 1 | 1 | 0 | 1 | 1 | 1 | 1 | 0 | 1 | 5 | Morales-Ivorra, I., Narváez, J., Gómez-Vaquero, C., Moragues, C., Nolla, J. M., Narváez, J. A., & Marín-López, M. A. (2022). Assessment of inflammation in patients with rheumatoid arthritis using thermography and machine learning: A fast and automated technique. RMD Open, 8(2). https://doi.org/10.1136/rmdopen-2022-002458 |
| 35 | Myasoedova | Toward Individualized Prediction of Response to Methotrexate in Early Rheumatoid Arthritis: A Pharmacogenomics-Driven Machine Learning Approach | 2022 | Predict Treatment Response | Random Forest | 71 | 71 | 71 | 0 | 0 | 0 | 1 | 0 | 1 | 1 | 2 | 0 | 2 | 2 | 2 | 0 | 9 | Myasoedova, E., Athreya, A. P., Crowson, C. S., Davis, J. M., Warrington, K. J., Walchak, R. C., Carlson, E., Kalari, K. R., Bongartz, T., Tak, P. P., van Vollenhoven, R. F., Padyukov, L., Emery, P., Morgan, A., Wang, L., Weinshilboum, R. M., & Matteson, E. L. (2022). Toward Individualized Prediction of Response to Methotrexate in Early Rheumatoid Arthritis: A Pharmacogenomics-Driven Machine Learning Approach. Arthritis Care and Research, 74(6), 879–888. https://doi.org/10.1002/acr.24834 |
| 36 | Norgeot | Assessment of a Deep Learning Model Based on Electronic Health Record Data to Forecast Clinical Outcomes in Patients With Rheumatoid Arthritis | 2019 | Score Disease Activity | NN | NA | 82 | NA | 0 | 0 | 0 | 0 | 0 | 0 | 2 | 2 | 1 | 1 | 2 | 0 | 1 | 9 | Norgeot, B., Glicksberg, B. S., Trupin, L., Lituiev, D., Gianfrancesco, M., Oskotsky, B., Schmajuk, G., Yazdany, J., & Butte, A. J. (2019). Assessment of a Deep Learning Model Based on Electronic Health Record Data to Forecast Clinical Outcomes in Patients With Rheumatoid Arthritis. JAMA Network Open, 2(3), e190606. https://doi.org/10.1001/jamanetworkopen.2019.0606 |
| 37 | Pauk | A computational method to differentiate rheumatoid arthritis patients using thermography data | 2022 | Improve Diagnostic Accuracy | NN | 84 | NA | NA | 0 | 0 | 0 | 0 | 0 | 0 | 1 | 1 | 0 | 0 | 1 | 0 | 0 | 3 | Pauk, J., Trinkunas, J., Puronaite, R., Ihnatouski, M., & Wasilewska, A. (2022). A computational method to differentiate rheumatoid arthritis patients using thermography data. Technology and Health Care, 30(1), 209–216. https://doi.org/10.3233/THC-219004 |

| | | | | | | | | | | | | | | | | | | | | | | | |
|---|---|---|---|---|---|---|---|---|---|---|---|---|---|---|---|---|---|---|---|---|---|---|---|
| 38 | Prasad | ATRPred: A machine learning based tool for clinical decision making of anti-TNF treatment in rheumatoid arthritis patients | 2022 | Predict Treatment Response | KNN | 76 | NA | NA | 0 | 0 | 0 | 1 | 0 | 1 | 2 | 1 | 0 | 1 | 1 | 0 | 2 | 7 | Prasad, B., McGeough, C., Eakin, A., Ahmed, T., Small, D., Gardiner, P., Pendleton, A., Wright, G., Bjourson, A. J., Gibson, D. S., & Shukla, P. (2022). ATRPred: A machine learning based tool for clinical decision making of anti-TNF treatment in rheumatoid arthritis patients. PLoS Computational Biology, 18(7). https://doi.org/10.1371/journal.pcbi.1010204 |
| 39 | Radke | Adaptive IoU Thresholding for Improving Small Object Detection: A Proof-of-Concept Study of Hand Erosions Classification of Patients with Rheumatic Arthritis on X-ray Images | 2023 | Assess Joint Damage | NN | 67 | 65 | 75 | 0 | 0 | 0 | 0 | 0 | 0 | 2 | 1 | 0 | 1 | 1 | 0 | 1 | 6 | Radke, K. L., Kors, M., Müller-Lutz, A., Frenken, M., Wilms, L. M., Baraliakos, X., Wittsack, H.-J., Distler, J. H. W., Abrar, D. B., Antoch, G., & Sewerin, P. (2023). Adaptive IoU Thresholding for Improving Small Object Detection: A Proof-of-Concept Study of Hand Erosions Classification of Patients with Rheumatic Arthritis on X-ray Images. Diagnostics, 13(1). https://doi.org/10.3390/diagnostics13010104 |
| 40 | Reed | Pilot study of a machine-learning tool to assist in the diagnosis of hand arthritis | 2022 | Improve Diagnostic Accuracy | NN Random Forest, Regression, SVM | 68 | NA | NA | 0 | 0 | 0 | 0 | 0 | 0 | 1 | 1 | 0 | 0 | 0 | 1 | 0 | 3 | Reed, M., Le Souëf, T., & Rampono, E. (2022). Pilot study of a machine-learning tool to assist in the diagnosis of hand arthritis. Internal Medicine Journal, 52(6), 959–967. https://doi.org/10.1111/imj.15173 |
| 41 | Srinivasan | A Framework of Faster CRNN and VGG16-Enhanced Region Proposal Network for Detection and Grade Classification of Knee RA | 2023 | Assess Joint Damage | NN | 60 | NA | NA | 0 | 0 | 0 | 0 | 0 | 0 | 1 | 1 | 0 | 0 | 1 | 0 | 0 | 3 | Srinivasan, S., Gunasekaran, S., Mathivanan, S. K., Jayagopal, P., Khan, M. A., Alasiry, A., Marzougui, M., & Masood, A. (2023). A Framework of Faster CRNN and VGG16-Enhanced Region Proposal Network for Detection and Grade Classification of Knee RA. Diagnostics, 13(8). https://doi.org/10.3390/diagnostics13081385 |
| 42 | Tao | Multiomics and Machine Learning Accurately Predict Clinical Response to Adalimumab and Etanercept Therapy in Patients With Rheumatoid Arthritis | 2021 | Predict Treatment Response | Random Forest | 70 | NA | NA | 0 | 0 | 0 | 1 | 0 | 1 | 2 | 2 | 0 | 1 | 2 | 1 | 1 | 9 | Tao, W., Concepcion, A. N., Vianen, M., Marijnissen, A. C. A., Lafeber, F. P. G. J., Radstake, T. R. D. J., & Pandit, A. (2021). Multiomics and Machine Learning Accurately Predict Clinical Response to Adalimumab and Etanercept Therapy in Patients With Rheumatoid Arthritis. Arthritis and Rheumatology, 73(2), 212–222. https://doi.org/10.1002/art.41516 |
| 43 | Venerito | A Machine Learning Approach for Predicting Sustained Remission in Rheumatoid Arthritis Patients on Biologic Agents | 2022 | Predict Treatment Response | Regression, Random Forest, Boosted Tree, KNN | 88 | NA | NA | 0 | 0 | 0 | 0 | 0 | 0 | 1 | 1 | 2 | 2 | 2 | 1 | 0 | 9 | Venerito, V., Angelini, O., Fornaro, M., Cacciapaglia, F., Lopalco, G., & Iannone, F. (2022). A Machine Learning Approach for Predicting Sustained Remission in Rheumatoid Arthritis Patients on Biologic Agents. Journal of Clinical Rheumatology, 28(2), E334–E339. https://doi.org/10.1097/RHU.0000000000001720 |
| 44 | Vodencarevic | Advanced machine learning for predicting individual risk of flares in rheumatoid arthritis patients tapering biologic drugs | 2021 | Predict Treatment Response | Regression, KNN, Naïve Bayes, Random Forest | 59 | NA | NA | 0 | 0 | 0 | 0 | 0 | 0 | 1 | 1 | 0 | 2 | 2 | 2 | 1 | 9 | Vodencarevic, A., Tascilar, K., Hartmann, F., Reiser, M., Hueber, A. J., Haschka, J., Bayat, S., Meinderink, T., Knitza, J., Mendez, L., Hagen, M., Krönke, G., Rech, J., Manger, B., Kleyer, A., Zimmermann-Rittereiser, M., Schett, G., & Simon, D. (2021). Advanced machine learning for predicting individual risk of flares in rheumatoid arthritis patients tapering biologic drugs. Arthritis Research and Therapy, 23(1). https://doi.org/10.1186/s13075-021-02439-5 |
| 45 | Zhou | RATING: Medical knowledge-guided rheumatoid arthritis assessment from multimodal ultrasound images via deep learning | 2022 | Score Disease Activity | NN | 70 | 70 | 69 | 0 | 0 | 0 | 0 | 0 | 0 | 2 | 2 | 0 | 1 | 1 | 2 | 2 | 10 | Zhou, Z., Zhao, C., Qiao, H., Wang, M., Guo, Y., Wang, Q., Zhang, R., Wu, H., Dong, F., Qi, Z., Li, J., Tian, X., Zeng, X., Jiang, Y., Xu, F., Dai, Q., & Yang, M. (2022). RATING: Medical knowledge-guided rheumatoid arthritis assessment from multimodal ultrasound images via deep learning. Patterns, 3(10). https://doi.org/10.1016/J.PATTER.2022.100592 |
| 46 | Plant | Profiling of Gene Expression Biomarkers as a Classifier of Methotrexate Nonresponse in Patients With Rheumatoid Arthritis | 2019 | Predict Treatment Response | Regression, Random Forest, Other | 77 | NA | NA | 0 | 0 | 0 | 0 | 0 | 0 | 1 | 1 | 1 | 1 | 1 | 1 | 1 | 7 | Plant, D., Maciejewski, M., Smith, S., Nair, N., Hyrich, K., Ziemek, D., Barton, A., & Verstappen, S. (2019). Profiling of Gene Expression Biomarkers as a Classifier of Methotrexate Nonresponse in Patients With Rheumatoid Arthritis. Arthritis & Rheumatology (Hoboken, N.J.), 71(5), 678–684. https://doi.org/10.1002/ART.40810 |
| 47 | Wu | Metagenomics Biomarkers Selected for Prediction of Three Different Diseases in Chinese Population | 2018 | Score Disease Activity | KNN, Regression, Random Forest, SVM, Boosted | 51 | NA | NA | 0 | 0 | 0 | 0 | 0 | 0 | 1 | 1 | 0 | 1 | 0 | 1 | 1 | 5 | Wu, H., Cai, L., Li, D., Wang, X., Zhao, S., Zou, F., & Zhou, K. (2018). Metagenomics Biomarkers Selected for Prediction of Three Different Diseases in Chinese Population. BioMed Research International, 2018. https://doi.org/10.1155/2018/2936257 |
| 48 | Chen | The prognostic value of whole-genome DNA methylation in response to Leflunomide in patients with Rheumatoid Arthritis | 2023 | Predict Treatment Response | Regression, Random Forest, Boosted Tree, SVM, Naïve Bayes | 77 | NA | NA | 0 | 0 | 0 | 1 | 0 | 1 | 2 | 1 | 1 | 0 | 0 | 1 | 1 | 6 | Chen, Y., Wang, Q., Liu, H., Jin, L., Feng, X., Dai, B., Chen, M., Xin, F., Wei, T., Bai, B., Fan, Z., Li, J., Yao, Y., Liao, R., Zhang, J., Jin, X., & Fu, L. (2023). The prognostic value of whole-genome DNA methylation in response to Leflunomide in patients with Rheumatoid Arthritis. Frontiers in Immunology, 14. https://doi.org/10.3389/fimmu.2023.1173187 |
| 49 | Okita | Automatic evaluation of atlantoaxial subluxation in rheumatoid arthritis by a deep learning model | 2023 | Assess Joint Damage | NN | 78 | NA | NA | 0 | 0 | 0 | 0 | 0 | 0 | 0 | 1 | 0 | 1 | 1 | 0 | 2 | 5 | Okita, Y., Hirano, T., Wang, B., Nakashima, Y., Minoda, S., Nagahara, H., & Kumanogoh, A. (2023). Automatic evaluation of atlantoaxial subluxation in rheumatoid arthritis by a deep learning model. Arthritis Research and Therapy, 25(1). https://doi.org/10.1186/s13075-023-03172-x |
| 50 | Rao | Machine Learning Approaches to Classify Self-Reported Rheumatoid Arthritis Health Scores Using Activity Tracker Data: Longitudinal Observational Study | 2023 | Score Disease Activity | Random Forest, Other | 92 | NA | NA | 0 | 0 | 0 | 0 | 0 | 0 | 1 | 0 | 0 | 1 | 1 | 1 | 1 | 5 | Rao, K., Speier, W., Meng, Y., Wang, J., Ramesh, N., Xie, F., Su, Y., Nowell, W. B., Curtis, J. R., & Arnold, C. (2023). Machine Learning Approaches to Classify Self-Reported Rheumatoid Arthritis Health Scores Using Activity Tracker Data: Longitudinal Observational Study. JMIR Formative Research, 7. https://doi.org/10.2196/43107 |

| | | | | | | | | | | | | | | | | | | | | | | | |
|---|---|---|---|---|---|---|---|---|---|---|---|---|---|---|---|---|---|---|---|---|---|---|---|
| 51 | Rothe | Fluorescence optical imaging feature selection with machine learning for differential diagnosis of selected rheumatic diseases | 2023 | Improve Diagnostic Accuracy | Boosted Tree | 76 | NA | NA | 0 | 0 | 0 | 0 | 0 | 0 | 2 | 1 | 0 | 1 | 0 | 2 | 1 | 7 | Rothe, F., Berger, J., Welker, P., Fiebelkorn, R., Kupper, S., Kiesel, D., Gedat, E., & Ohrndorf, S. (2023). Fluorescence optical imaging feature selection with machine learning for differential diagnosis of selected rheumatic diseases. Frontiers in Medicine, 10. https://doi.org/10.3389/fmed.2023.1228832 |
| 52 | Saleh | USE OF SOME BONE-RELATED CYTOKINES AS PREDICTORS FOR RHEUMATOID ARTHRITIS SEVERITY BY NEURAL NETWORK ANALYSIS | 2023 | Score Disease Activity | NN | 57 | NA | NA | 0 | 0 | 1 | 0 | 0 | 1 | 1 | 1 | 0 | 0 | 1 | 0 | 0 | 3 | Saleh, R. O., Mahmood, L. A., Mohammed, M. A., Al-Rawi, K. F., & Al-Hakeim, H. K. (2023). USE OF SOME BONE-RELATED CYTOKINES AS PREDICTORS FOR RHEUMATOID ARTHRITIS SEVERITY BY NEURAL NETWORK ANALYSIS. Russian Journal of Infection and Immunity, 13(1), 147–155. https://doi.org/10.15789/2220-7619-UOS-2008 |
